# Using Social Determinants of Health ICD-10 Z-codes to Identify Non-Medical Factors among Asthma Hospitalizations in the United States, 2016–2022

**DOI:** 10.64898/2026.08.19.26360844

**Authors:** Nianyang Wang, Huang Huang, Jun Chu, Joy Hsu

**Affiliations:** Centers for Disease Control and Prevention, National Center for Environmental Health 4770 Buford Hwy, Atlanta, GA 30341; Augusta University, School of Public Health; University of Maryland Baltimore County, Department of Sociology, Anthropology, and Public Health; Centers for Disease Control and Prevention, National Center for Environmental Health

**Keywords:** asthma, social determinants of health, Z-codes, socioeconomic factors, hospital

## Abstract

**Objectives:** Healthcare data can reveal actionable opportunities to prevent asthma hospitalizations. Limited national-level data exist regarding social determinants of health (SDOH) and asthma hospitalizations. We examined SDOH-related International Classification of Diseases, Tenth Revision (ICD-10) Z-codes in national administrative data on asthma hospitalizations and described patient- and hospital-level characteristics associated with documented SDOH Z-codes.

**Methods:** Pooled cross-sectional analysis of 2016–2022 Nationwide Inpatient Sample for 200,452 U.S. hospitalizations (all ages) with a primary diagnosis of asthma. Presence of SDOH Z-codes (codes Z55– Z65) assessed by descriptive statistics and multivariable logistic regression to calculate odds ratios (ORs) and 95% confidence intervals (95% CIs) for associations between SDOH Z-codes and patient- and hospital-level characteristics.

**Results:** In unweighted analyses, 3,149 asthma hospitalizations had SDOH Z-codes (1.57%). The most common SDOH Z-codes were homelessness (Z59.0; n=942) and unemployment (Z56.0; n=349).

Weighted chi-square analyses found all selected variables were associated with asthma hospitalization SDOH Z-code documentation. Logistic regression results varied; adjusted odds for SDOH Z-code documentation were higher for asthma hospitalizations involving male patients (aOR=1.51; 95% CI, 1.39–1.63; *P* < .001) compared to female patients. Asthma hospitalizations involving rural hospitals had lower odds of SDOH Z-codes documentation (aOR=0.57; 95% CI, 0.47–0.70; *P* < .001) compared to urban teaching hospitals.

**Conclusions:** National 2016–2022 data indicate housing- and employment-related Z-codes were the most commonly documented SDOH within asthma hospitalizations. Future analyses could consider establishing causality and exploring how relationships between these SDOH may be used by public health practitioners and others to improve program interventions.

## Introduction

Asthma affects 25 million Americans and is associated with nearly 100,000 hospitalizations annually, many of which are preventable.^1,2^ While asthma hospitalizations are strongly associated with exposures such as tobacco smoke and respiratory viruses, some data suggest they are also associated with social determinants of health (SDOH).^3–6^ These SDOH are nonmedical factors that can affect health outcomes where people are born, live, learn, work, play, worship, and age such as safe housing, education, and job opportunities.^7,8^ Addressing SDOH (e.g., housing and employment related issues) can improve health and potentially prevent some asthma hospitalizations. However, little is known about documented SDOH among patients with asthma hospitalizations.^3–6^

Routine review of specific healthcare data, including SDOH data, can help practitioners identify actionable opportunities to prevent hospitalizations. For example, hospitalization data linked to low access to SDOH could potentially help practitioners and their partners (e.g., public health agencies, community organizations) identify people who could benefit from social services. Since 2016, the International Classification of Diseases, Tenth Revision, Clinical Modification (ICD-10-CM) has included SDOH Z-codes that indicate homelessness, unemployment, and other nonmedical factors relevant to health outcomes.^9^ Research has indicated SDOH Z-code documentation is infrequent (<2% of all-cause hospitalizations in one study^10^) and may be underutilized. However, SDOH Z-code analyses can suggest opportunities to help some people experiencing hospitalizations that could be avoided with greater access to SDOH. For example, a large Medicare data analysis (not asthma-specific) indicated homelessness was the most commonly documented SDOH Z-code.^11^ Other analyses have demonstrated associations between SDOH Z-code documentation and hospital readmission risk.^11–13^

While some data suggest relationships between access to SDOH and asthma prevalence,^2–5^ no national studies have examined SDOH Z-codes among asthma hospitalizations. Because SDOH Z-codes may identify actionable opportunities to help people with asthma, we sought to advance Truong et al.’s work on all-cause hospitalizations and SDOH Z-codes.^10^ Our first objective was to analyze and describe documented SDOH Z-codes in asthma hospitalization data. Our second objective was to assess these codes’ relationships to selected patient- and hospital-level characteristics.

## Methods

We conducted a pooled cross-sectional analysis of secondary data from the 2016–2022 National Inpatient Sample (NIS). The NIS is part of the Healthcare Cost and Utilization Project (HCUP) administered by the Agency for Healthcare Research and Quality. NIS data provide a nationally representative annual sample of roughly 20% of all U.S. hospitalizations, with data on patient diagnoses, sociodemographic factors, and hospital characteristics. The data do not contain personal identifiers, and we adhered to the NIS Data Use Agreement.^14^ This analysis was determined to be exempt from human research protections review by Augusta University, the University of Maryland, and CDC.

Our analysis focused on asthma hospitalizations, defined by a primary diagnosis of asthma (ICD- 10 code J45).^15^ The primary outcome was whether the asthma hospitalization documented any SDOH ICD-10 Z-codes (ICD-10 codes Z55–Z65). These codes are assigned by healthcare providers during care delivery or by medical coders reviewing patients’ medical records. Our independent variables comprised seven sociodemographic and hospital factors as studied and identified in Truong et al’s research to be associated with all-cause admissions:^10^ age, sex, race/ethnicity, primary payer, estimated household income quartile based on patient’s ZIP code, hospital region, and hospital category (urban teaching, urban non-teaching, or rural). We excluded hospitalizations missing any of these factors. Similar to prior publications,^16,17^ we accounted for NIS’s complex survey design by using NIS-provided sample weights. We also divided by seven for the primary 2016–2022 analysis (and by four for the 2016–2019 sub-analysis). Our final sample included 200,452 asthma hospitalizations.^16–18^

We first analyzed unweighted data to assess SDOH Z-code distribution among asthma hospitalizations.^14^ Next, we used weighted data to generate descriptive statistics of asthma hospitalizations stratified by the presence of SDOH Z-codes. We performed chi-square tests to determine between-group differences. Lastly, we used the same variables as Truong et al^10^ to create univariable and multivariable logistic regressions examining the association between seven selected independent variables and asthma hospitalization. We calculated unadjusted and adjusted odds ratios (OR and aOR) and 95% confidence intervals (CIs).

We conducted a robustness check of re-estimating our multivariable regression while excluding the years 2020–2022 to eliminate potential impacts of the COVID-19 pandemic (eTable 1 in the Supplement).^19^ We used Stata version 18 (StataCorp) to conduct all analyses; statistical significance was set at α=.05.

## Results

In unweighted analyses, our sample was 200,452 asthma hospitalizations during 2016–2022. Of these, 3,149 (1.57%) had SDOH Z-codes. A subset had two or more associated SDOH Z-codes (n=215; 6.83% of all asthma hospitalizations with SDOH Z-codes). Table 1 shows the distribution of SDOH-related Z-codes among asthma hospitalizations. “Problems related to housing and economic circumstances” was the most prevalent Z-code category (Z59; n=1,702; 49.87%). It was followed by “Problems related to upbringing” (Z62; n=415; 12.16%) and “Problems related to employment and unemployment” (Z56; n=359; 10.52%). By individual Z-code, the most documented SDOH Z-codes were “Homelessness” (Z59.0; n=942) and “Unemployment” (Z56.0; n=349).

**Table 1.** Unweighted Frequencies of Social Determinants of Health-Related Z-codes Among Asthma Hospitalizations with These Z-codes, by Z-code Category — National Inpatient Sample, 2016–2022.

| **SDOH Z-code and Description** | **Z-code**<br>**Counts** | **Z-code**<br>**Category**<br>**Counts (%)**<br>**(N=3,413<sup>1</sup>)** |
| --- | --- | --- |
| **Z55 - Problems related to education and literacy** |  | 22 (0.64%) |
| Z55.0 Illiteracy and low-level literacy | <11<sup>2</sup> |  |
| Z55.3 Underachievement in school | <11 |  |
| Z55.5 Less than a high school diploma | <11 |  |
| Z55.8 Other problems related to education and literacy | <11 |  |
| Z55.9 Problems related to education and literacy, unspecified | <11 |  |
| **Z56 - Problems related to employment and unemployment** |  | 359 (10.52%) |
| Z56.0 Unemployment, unspecified | 349 |  |
| Z56.1 Change of job | <11 |  |
| Z56.2 Threat of job loss | <11 |  |
| Z56.3 Stressful work schedule | <11 |  |
| Z56.5 Uncongenial work environment | <11 |  |
| Z56.6 Other physical and mental strain related to work | <11 |  |
| Z56.89 Other problems related to employment | <11 |  |
| Z56.9 Unspecified problems related to employment | <11 |  |
| **Z57 - Occupational exposure to risk factors** |  | 206 (6.04%) |
| Z57.2 Occupational exposure to dust | 71 |  |
| Z57.31 Occupational exposure to environmental tobacco smoke | 41 |  |
| Z57.39 Occupational exposure to other air contaminants | 18 |  |
| Z57.4 Occupational exposure to toxic agents in agriculture | <11 |  |
| Z57.5 Occupational exposure to toxic agents in other industries | 53 |  |
| Z57.6 Occupational exposure to extreme temperature | <11 |  |
| Z57.8 Occupational exposure to other risk factors | <11 |  |
| Z57.9 Occupational exposure to unspecified risk factor | <11 |  |
| **Z59 - Problems related to housing and economic circumstances** |  | 1,702 (49.87%) |
| Z59.0 Homelessness | 942 |  |
| Z59.00 Homelessness unspecified | 109 |  |
| Z59.01 Sheltered homelessness | 68 |  |
| Z59.02 Unsheltered homelessness | 23 |  |
| Z59.1 Inadequate Housing | 16 |  |
| Z59.3 Problems related to living in residential institution | <11 |  |
| Z59.4 Lack of adequate food | 14 |  |
| Z59.41 Food insecurity | 37 |  |
| Z59.48 Other specified lack of adequate food | <11 |  |
| Z59.5 Extreme poverty | <11 |  |
| Z59.6 Low income | 82 |  |
| Z59.7 Insufficient social insurance and welfare support | 161 |  |
| Z59.8 Other problems related to housing and economic circumstances | 86 |  |
| Z59.812 Housing instability, housed, homelessness in past 12 months | <11 |  |
| Z59.819 Housing instability, housed unspecified | 12 |  |
| Z59.82 Transportation insecurity | <11 |  |
| Z59.86 Financial insecurity | <11 |  |
| Z59.89 Other problems related to housing and economic circumstances | 28 |  |
| Z59.9 Problem related to housing and economic circumstances, unspecified | 110 |  |
| **Z60 - Problems related to social environment** |  | 315 (9.23%) |
| Z60.0 Problems of adjustment to life-cycle transitions | <11 |  |
| Z60.2 Problems related to living alone | 188 |  |
| Z60.3 Acculturation difficulty | 14 |  |
| Z60.4 Social exclusion and rejection | 11 |  |
| Z60.5 Target of (perceived) adverse discrimination and persecution | <11 |  |
| Z60.8 Other problems related to social environment | 47 |  |
| Z60.9 Problem related to social environment, unspecified | 53 |  |
| **Z62 - Problems related to upbringing** |  | 415 (12.16%) |
| Z62.0 Inadequate parental supervision and control | <11 |  |
| Z62.21 Child in welfare custody | 203 |  |
| Z62.29 Other upbringing away from parents | <11 |  |
| Z62.810 Personal history of abuse in childhood | 143 |  |
| Z62.811 Personal history of psychological abuse in childhood | <11 |  |
| Z62.812 Personal history of neglect in childhood | 14 |  |
| Z62.819 Personal history of unspecified abuse in childhood | 19 |  |
| Z62.820 Parent-child conflict | <11 |  |
| Z62.822 Parent-foster child conflict | <11 |  |
| Z62.890 Parent-child estrangement NEC (Not Elsewhere Classifiable) | <11 |  |
| Z62.898 Other specified problems related to upbringing | <11 |  |
| Z62.9 Problem related to upbringing, unspecified | <11 |  |
| **Z63 - Other problems related to primary support group, including family circumstances** |  | 302 (8.85%) |
| Z63.0 Problems in relationship with spouse or partner | 12 |  |
| Z63.32 Other absence of family member | <11 |  |
| Z63.4 Disappearance and death of family member | 106 |  |
| Z63.5 Disruption of family by separation and divorce | 44 |  |
| Z63.6 Dependent relative needing care at home | <11 |  |
| Z63.72 Alcoholism and drug addiction in family | <11 |  |
| Z63.79 Other stressful life events affecting family and household | 37 |  |
| Z63.8 Other specified problems related to primary support group | 71 |  |
| Z63.9 Problem related to primary support group, unspecified | 19 |  |
| **Z65 - Problems related to other psychosocial circumstances** |  | 92 (2.70%) |
| Z65.1 Imprisonment and other incarceration | 36 |  |
| Z65.3 Problems related to other legal circumstances | 22 |  |
| Z65.5 Exposure to disaster, war, and other hostilities | <11 |  |
| Z65.8 Other specified problems related to psychosocial circumstances | 19 |  |
| Z65.9 Problem related to unspecified psychosocial circumstances | 14 |  |

SDOH: social determinants of health.
Data: 2016–2022 National Inpatient Sample (NIS).
<sup>1</sup> The total number of asthma hospitalizations with SDOH Z-codes is 3,149 and some hospitalizations had more than one SDOH Z-code.
<sup>2</sup>Numbers less than 11 are suppressed due to the NIS data use agreement.

Next, we used weighted data to conduct chi-square analyses and found that the prevalence of documented SDOH Z-codes in asthma hospitalizations significantly differed among categories of each factors we studied (Table 2). Univariable and multivariable logistic regression results (Table 3) revealed more details about patient- and hospital-level factors associated with SDOH Z-code documentation.

**Table 2.**
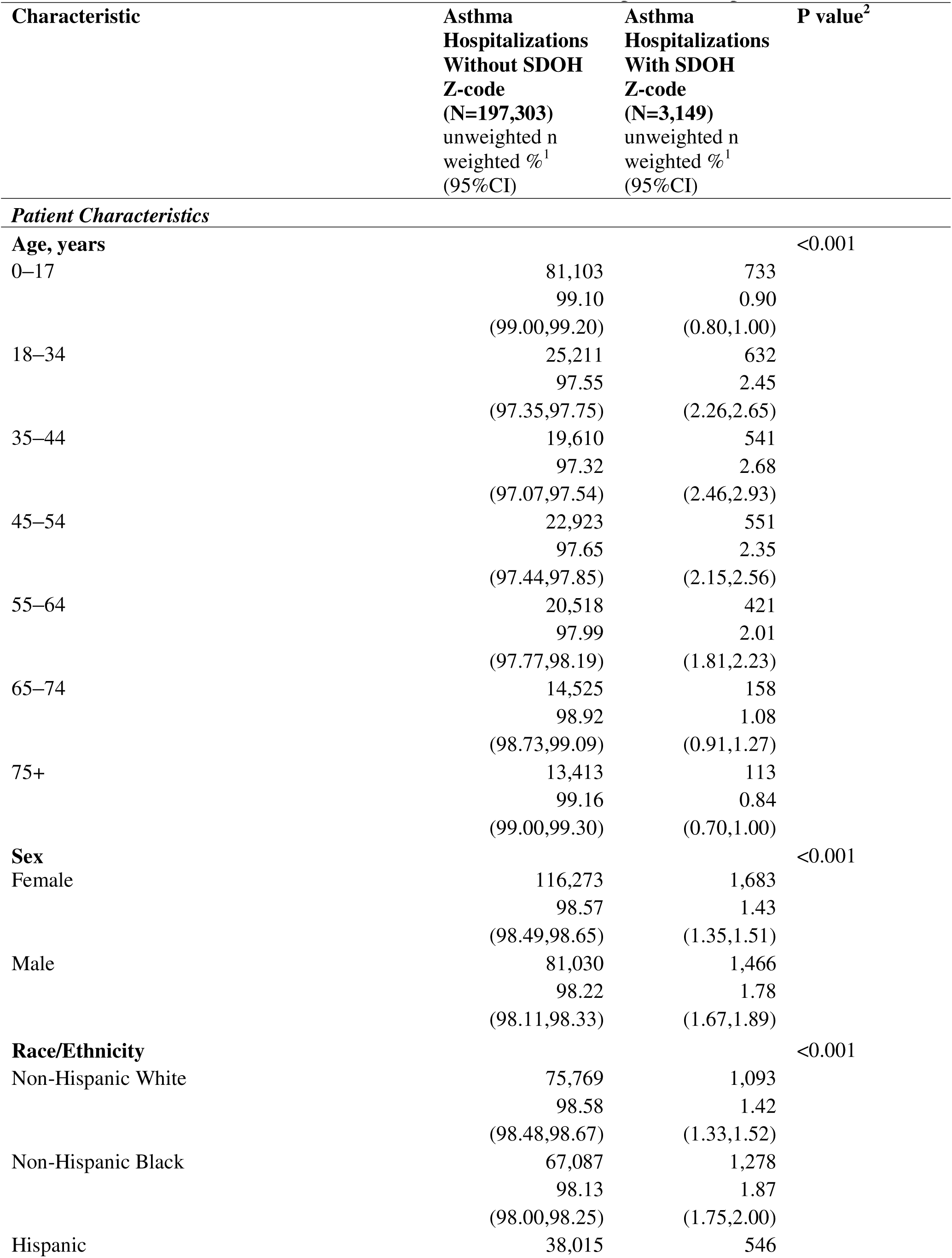
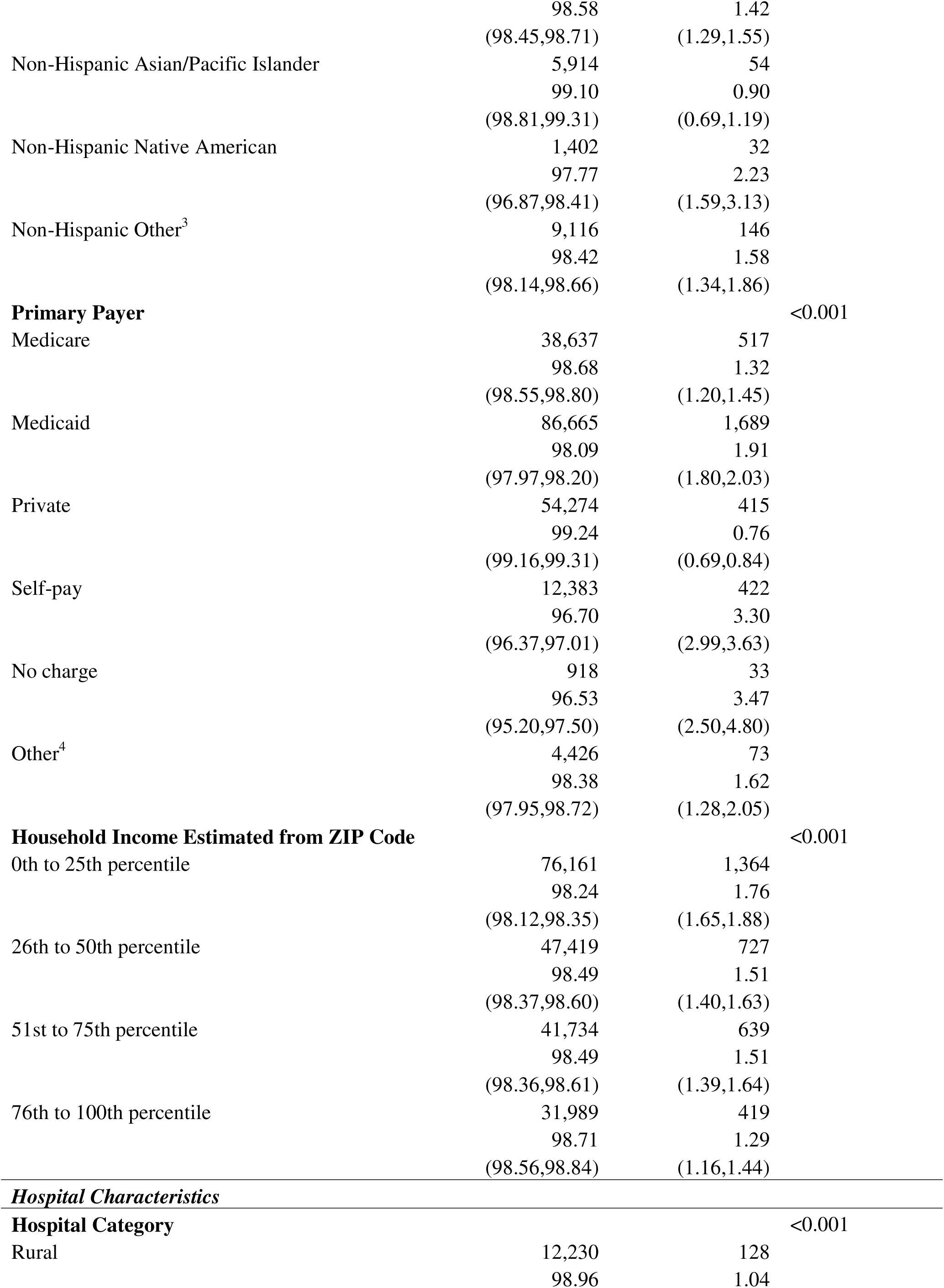
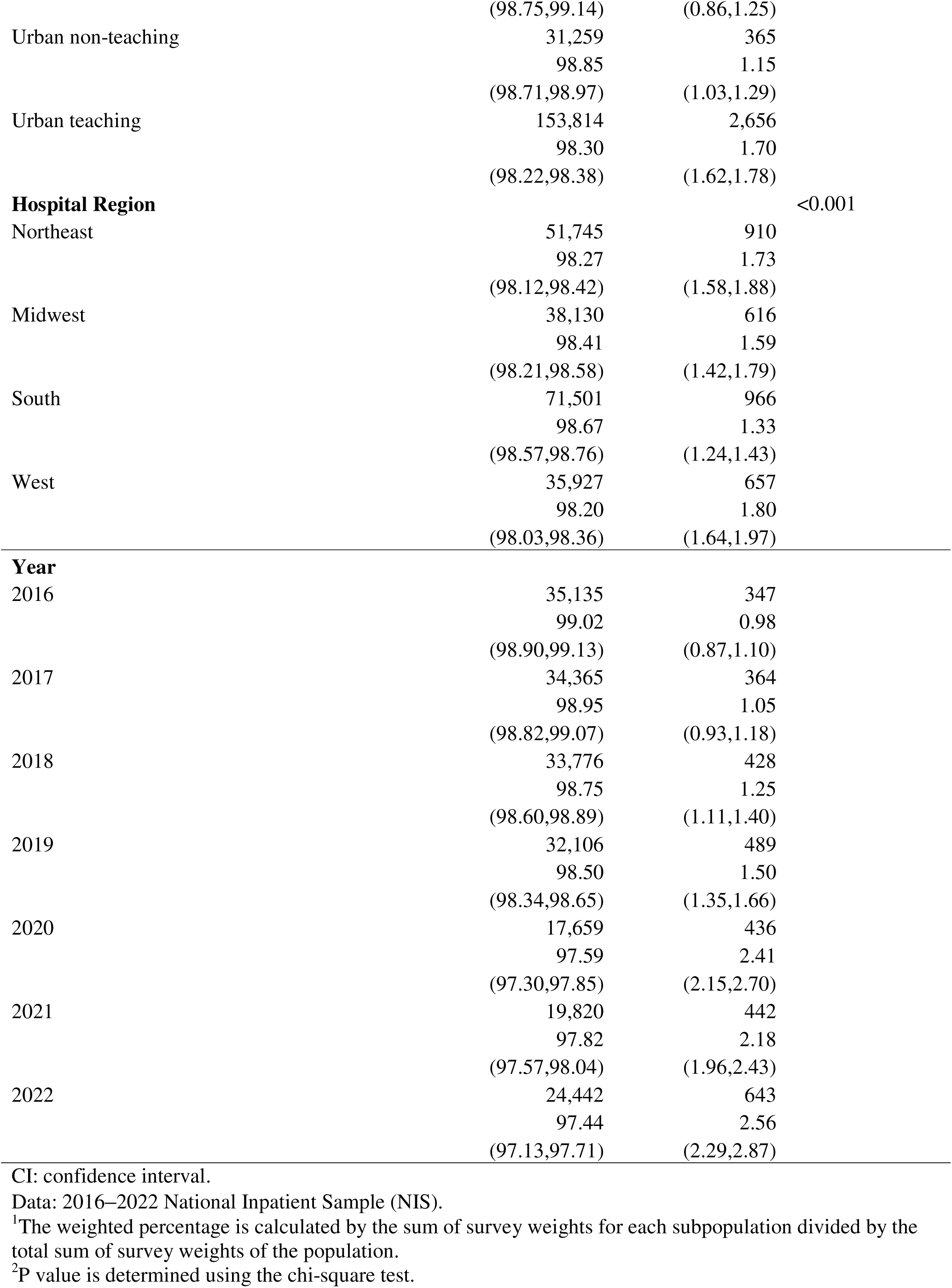
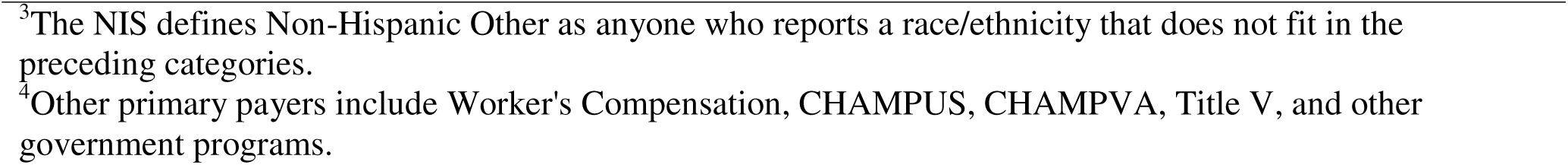
Sample Characteristics of Asthma Hospitalization patients in the United States Stratified by Presence of Social Determinants of Health Z-codes — National Inpatient Sample, 2016–2022.

**Table 3.**
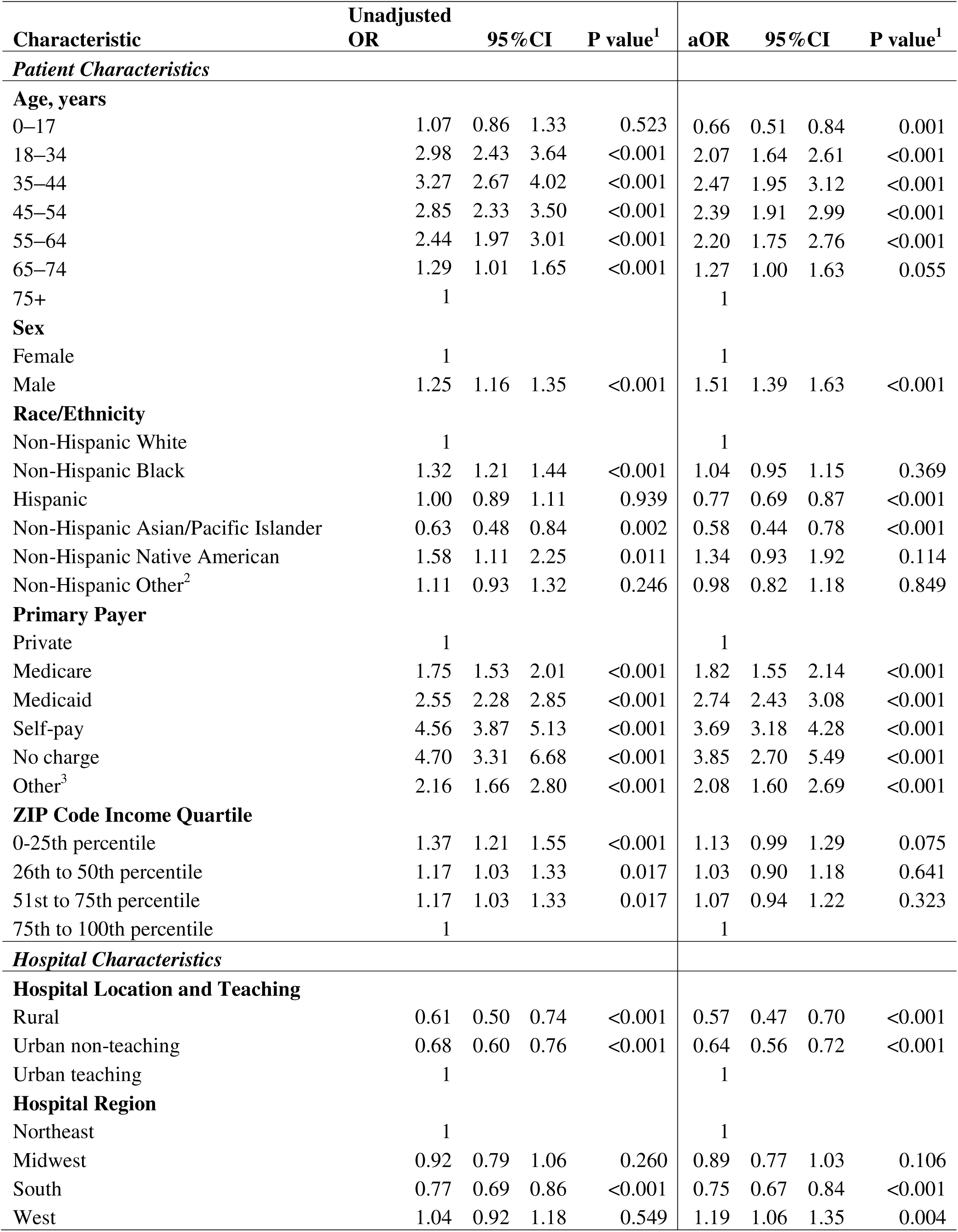
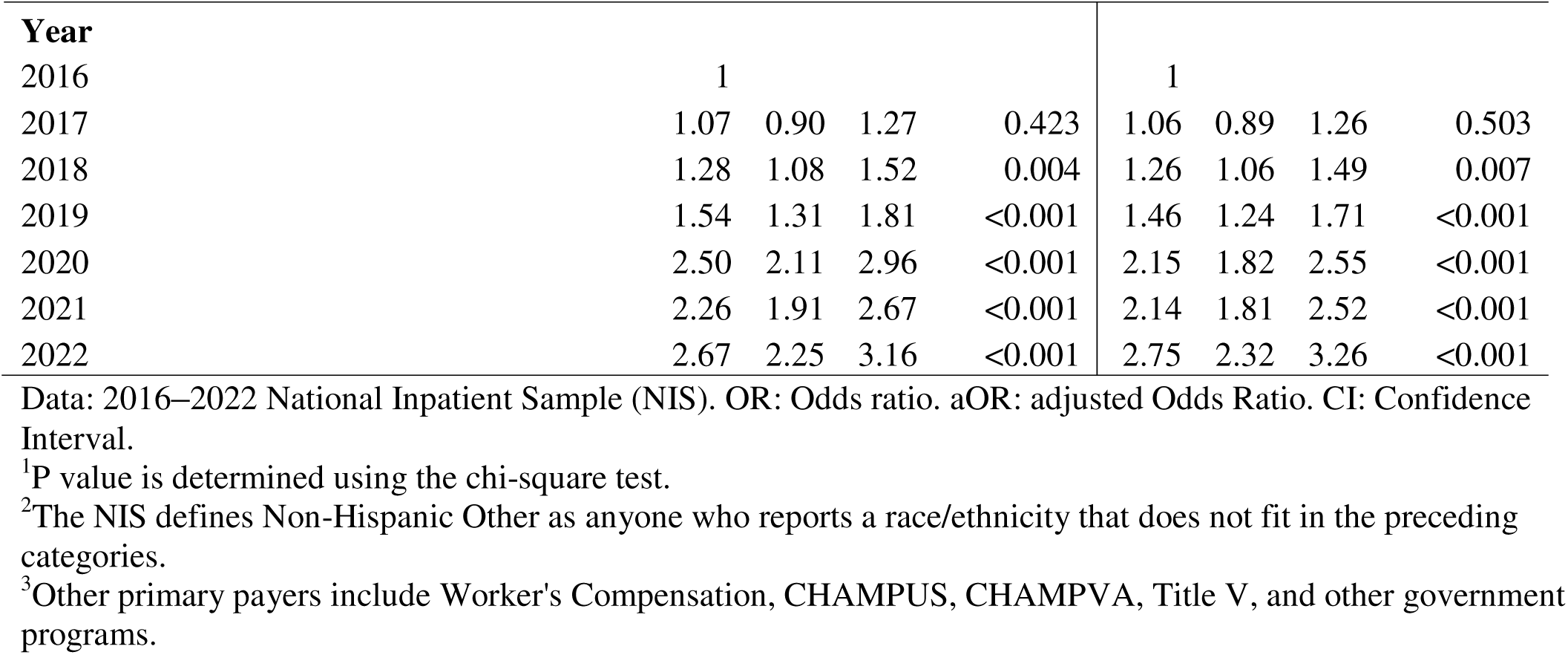
Univariable and Multivariable Logistic Regression Results of Documented Social Determinants of Health Z-codes Among Asthma Hospitalizations — National Inpatient Sample, 2016–2022.

Compared to the reference group of asthma hospitalizations involving patients ages 75+ years, those involving patients ages 18–34 years (aOR=2.07; 95% CI, 1.64–2.61; *P* < .001), ages 35–44 years (aOR=2.47; 95% CI, 1.95–3.12; *P* < .001), ages 45–54 years (aOR=2.39; 95% CI, 1.91–2.99; *P* < .001), and ages 55-64 (aOR=2.20; 95% CI, 1.75–2.76; *P* < .001) had higher odds of SDOH Z-code documentation in both unadjusted and adjusted analyses. Asthma hospitalizations involving patients ages 65–74 years had higher odds of SDOH documentation compared to the reference group in unadjusted analyses (OR=1.29; 95% CI, 1.01–1.65; *P* < .001), but the association was not statistically significant in adjusted analyses (aOR=1.27; 95% CI, 1.00–1.63; *P* = .055). Interestingly, asthma hospitalizations involving patients ages 0–17 years had lower odds of SDOH documentation compared to the reference group in adjusted analyses (aOR=0.66; 95% CI, 0.51–0.84; *P* = .001), but the association was not statistically significant in unadjusted analyses (OR=1.07; 95% CI, 0.86–1.33; *P* < .001).

In terms of other patient demographics, asthma hospitalizations involving males had higher unadjusted and adjusted odds of SDOH Z-code documentation (aOR=1.51; 95% CI, 1.39–1.63; *P* < .001) compared to those involving females. Compared to asthma hospitalizations involving non-Hispanic White patients, those involving non-Hispanic Native American patients had higher unadjusted odds of SDOH Z-code documentation (OR=1.58; 95% CI, 1.11–2.25; *P* = .011), but this relationship was not significant in adjusted analyses (aOR=1.34; 95% CI, 0.93–1.92; *P* = .114). Asthma hospitalizations involving non-Hispanic Asian and Pacific Islander patients had lower unadjusted and adjusted odds of SDOH Z-code documentation (aOR=0.54; 95% CI, 0.39–0.75; *P* < .001) than the reference group. Asthma hospitalizations involving non-Hispanic Black patients had higher odds of SDOH documentation than the reference group in unadjusted analyses (OR=1.32; 95% CI, 1.21–1.44; *P* < .001), but the association was not statistically significant in adjusted analyses (aOR=1.04; 95% CI, 0.95–1.15; *P* = .369). Conversely, asthma hospitalizations involving Hispanic patients had lower odds of SDOH documentation than the reference group in adjusted analyses (aOR=0.77; 95% CI, 0.69–0.87; *P* < .001), but the association was not statistically significant in unadjusted analyses (OR=1.00; 95% CI, 0.89–1.11; *P* = .939).

Asthma hospitalizations had higher adjusted odds of SDOH Z-code documentation if the patient’s insurance status was Medicare (aOR=1.82; 95% CI, 1.55–2.14; *P* < .001), Medicaid (aOR=2.74; 95% CI, 2.43–3.08; *P* < .001), self-pay (aOR=3.69; 95% CI, 3.18–4.28; *P* < .001), no charge (aOR=3.85; 95% CI, 2.70–5.49; *P* < .001), or other insurance (aOR=2.08; 95% CI, 1.60–2.69; *P* < .001) compared to private insurance. We found no statistically significant associations between income and SDOH Z-code documentation in multivariable analyses adjusted for covariates, including payer type.

Adjusted ORs were lower among asthma hospitalizations in rural hospitals (aOR=0.57; 95% CI, 0.47–0.70; *P* < .001) and urban non-teaching hospitals (aOR=0.64; 95% CI, 0.56–0.73; *P* < .001) compared to urban teaching hospitals. Regarding hospital region, asthma hospitalizations in the South (aOR=0.75; 95% CI, 0.67–0.84; *P* < .001) had lower adjusted odds of SDOH Z-code documentation compared to those in the Northeast, while hospitals in the West had higher adjusted odds (aOR=1.19; 95% CI, 1.06–1.35; *P* = .004). Lastly, from 2018 onwards, asthma hospitalizations had higher adjusted odds of SDOH Z-code documentation compared to 2016, whereas 2017-associated odds did not differ significantly compared to 2016.

Our robustness checks produced mostly consistent significance compared to our main results.

However, data restricted to 2016–2019 (see eTable 1 in the Supplement) showed hospitalizations involving non-Hispanic Other Race patients had lower adjusted odds of SDOH Z-code documentation (aOR=0.74; 95% CI, 0.56–0.96; *P* = .026) compared to hospitalizations involving non-Hispanic White patients, while the main results did not show this difference. In addition, asthma hospitalizations involving patients ages 0–17 years were not significant (aOR=0.93; 95% CI, 0.65–1.33; *P* = .684) compared to those involving patients ages 75+, while the main results showed lower significant odds.

## Discussion

This national study described individual and hospital-level factors associated with SDOH Z-code documentation and asthma hospitalizations. Our study illustrates how healthcare data can reveal actionable opportunities to prevent asthma hospitalizations. For example, our findings that housing- and employment-related Z-codes were the most common SDOH concerns documented in asthma hospitalizations add to literature regarding modifiable SDOH and asthma hospitalizations. Public health officials can consider using SDOH Z-codes to identify patients with recent asthma hospitalizations who might benefit from tailored support, such as local services for people experiencing homelessness or unemployment.

Our data analysis found 1.57% of all asthma hospitalizations had SDOH Z-codes in 2016–2022, which is comparable to the 1.9% reported by Truong et al.’s analysis of all-cause hospitalizations.^13^ Similarly, our finding that homelessness and unemployment were the most commonly documented SDOH Z-codes in asthma hospitalizations aligned with Truong et al.^10^ Our adjusted analysis results for each race/ethnicity category reflect those in Truong et al.^10^ However, our results for household income are not significant. Truong et al. showed significantly higher adjusted odds for patients with household incomes at the lowest and second lowest quartiles compared to the highest quartile.^10^

Our results also align with other publications. For example, a New York State study reported the likelihood of asthma hospitalization among children and adolescents was 31 times higher if they were experiencing homelessness.^21^ Another study found neighborhood unemployment rates were positively associated with an increased risk of asthma hospitalizations.^22^ We found asthma hospitalizations involving rural and urban non-teaching hospitals had lower adjusted odds of documenting SDOH Z-codes than those involving urban teaching hospitals. This parallels research showing a positive association between teaching hospitals and screening for health-related social needs.^23^ Also, 2022 data from the American Hospital Association Annual Survey indicated 83% of U.S. hospitals reported collecting social needs data, and hospital resource levels, including size, rurality, and critical access status, were associated with these data collection activities.^24^ Moreover, our national data complement a 2020 California study showing that screening for SDOH during asthma home health visits was associated with fewer hospitalization days.^25^

Nevertheless, not all our results were consistent with existing literature. For instance, one survey-based study indicated that Hispanic and non-Hispanic Black adults (with or without asthma) reported more unmet social needs than non-Hispanic White adults.^26^ However, our analysis of asthma-specific hospital administrative data indicated no differences or lower adjusted odds of documented SDOH Z-codes among these racial/ethnic groups. Also, we found no differences across household income quartiles in our analysis. In contrast, a previous survey showed 54% of U.S. adults (with or without asthma) living below the poverty level self-reported multiple unmet social needs.^27^ Possible reasons for the differences between our results and previous data include disparate data sources (i.e., survey versus administrative data) and study populations (i.e., people with asthma versus a broader population).

Strengths of our analysis include the use of recent national data. To our knowledge, this is the first national analysis of SDOH Z-codes in asthma hospitalizations. Our findings illustrate how administrative data, including SDOH Z-codes, can complement survey data. Together, they can potentially advance understanding of nonmedical factors relevant to asthma morbidity.

Our analysis has some limitations. First, we used a cross-sectional design and thus cannot infer causality. Also, collider bias is possible. Patients with lower access to SDOH may be more likely to be hospitalized, even if the documented SDOH factor is not the cause for hospitalization.^28^ In addition, the NIS does not have data from all U.S. hospitals and may underrepresent specialty hospitals such as psychiatric hospitals. Furthermore, the data do not contain variables to study more granular geographic data (census tract) or community-level factors that have been associated with poorer asthma-related health outcomes.^29^ Our sample size was insufficient to stratify by region to assess if race/ethnicity associations with SDOH Z-codes changed by region. Additionally, asthma-related health outcomes among Hispanic and non-Hispanic Asian persons can be influenced by language barriers, which the currently available SDOH Z-codes do not examine.^30^ Moreover, our Medicare-related findings were surprisingly unlike our results involving people ages 75+ years, but this study was not designed to further investigate this observed phenomenon. Finally, the deidentified data do not allow us to follow individual patients over time to track hospital readmissions.

## Conclusion

Administrative data, including SDOH Z-codes, can advance understanding of factors relevant to asthma morbidity. These data can also reveal actionable opportunities for asthma public health programs, healthcare systems, community organizations and others interested in preventing asthma hospitalizations. For example, we found that housing- and employment-related Z-codes were the most common SDOH concerns documented in asthma hospitalizations. Addressing these issues could improve the health of people with asthma. Further analyses would allow for defining the degree of causality and exploring how public health practitioners and others could use SDOH Z-codes to support people with asthma.

## Data Availability

The data used in this study were obtained from the Healthcare Cost and Utilization Project (HCUP), sponsored by the Agency for Healthcare Research and Quality (AHRQ). Because HCUP data are subject to the HCUP Data Use Agreement, the authors are not permitted to redistribute the underlying data or make them publicly available.

## Acknowledgements

The authors thank W. Dana Flanders (Rollins School of Public Health) and Sara Jacenko (Centers for Disease Control and Prevention) for their contributions to this article. We thank the Agency for Healthcare Research and Quality (AHRQ) for providing the data. No copyrighted material, surveys, instruments, or tools were used in the research described in this article.

## Author Contributions

NW: Conceptualization, Data Curation, Formal Analysis, Investigation, Project administration, Resources, Software, Visualization, Writing - Original Draft, Review & Editing. HH: Methodology, Writing - Original Draft, Review & Editing. JC: Validation, Writing - Original Draft, Review & Editing. JH: Supervision, Writing - Original Draft, Review & Editing.

## Statements and Declarations

The findings and conclusions in this report are those of the authors and do not necessarily represent the official position of the Centers for Disease Control and Prevention or AHRQ.

## Ethical considerations

This analysis was determined to be exempt from human research protections review by Augusta University, the University of Maryland, and CDC.

## Consent to participate

Not applicable

## Consent for publication

Not applicable

