## Supplemental Table 1 for "Using Social Determinants of Health ICD-10 Z-codes to Identify Non-Medical Factors among Asthma Hospitalizations in the United States, 2016–2022"

Supplement eTable 1. Multivariable Logistic Regression of Documented Social Determinants	of Health Z-codes among Asthma Hospitalizations — National Inpatient Sample, 2016–2019

| **Characteristic** | **aOR** | **95%CI** | | **P value^1^** |
| --- | --- | --- | --- | --- |
| ***Patient Characteristics*** |  |  |  |  |
| **Age, years** |  |  |  |  |
| 0–17 | 0.93 | 0.65 | 1.33 | 0.684 |
| 18–34 | 2.98 | 1.11 | 4.21 | <0.001 |
| 35–44 | 3.53 | 2.50 | 5.00 | <0.001 |
| 45–54 | 3.31 | 2.36 | 4.65 | <0.001 |
| 55–64 | 2.74 | 1.94 | 3.85 | <0.001 |
| 65–74 | 1.44 | 0.99 | 2.10 | 0.055 |
| 75+ | 1 |  |  |  |
| **Sex** |  |  |  |  |
| Female | 1 |  |  |  |
| Male | 1.48 | 1.33 | 1.66 | <0.001 |
| **Race/Ethnicity** |  |  |  |  |
| Non-Hispanic White | 1 |  |  |  |
| Non-Hispanic Black | 1.02 | 0.89 | 1.15 | 0.815 |
| Hispanic | 0.70 | 0.59 | 0.82 | <0.001 |
| Non-Hispanic Asian/Pacific Islander | 0.57 | 0.39 | 0.85 | 0.005 |
| Non-Hispanic Native American | 2.12 | 1.41 | 3.19 | <0.001 |
| Non-Hispanic Other^2^ | 0.74 | 0.56 | 0.96 | 0.026 |
| **Primary Payer** |  |  |  |  |
| Private | 1 |  |  |  |
| Medicare | 2.06 | 1.68 | 2.54 | <0.001 |
| Medicaid | 3.13 | 2.65 | 3.68 | <0.001 |
| Self-pay | 3.86 | 3.14 | 4.73 | <0.001 |
| No charge | 4.22 | 2.63 | 6.76 | <0.001 |
| Other^3^ | 1.93 | 1.31 | 2.84 | 0.001 |
| **Household Income Estimated from ZIP Code** |  |  |  |  |
| 0th to 25th percentile | 1.00 | 0.84 | 1.20 | 0.998 |
| 26th to 50th percentile | 0.92 | 0.76 | 1.10 | 0.351 |
| 51st to 75th percentile | 0.99 | 0.82 | 1.19 | 0.903 |
| 75th to 100th percentile | 1 |  |  |  |
| ***Hospital Characteristics*** |  |  |  |  |
| **Hospital Category** |  |  |  |  |
| Rural | 0.57 | 0.44 | 0.73 | <0.001 |
| Urban non-teaching | 0.65 | 0.55 | 0.76 | <0.001 |
| Urban teaching | 1 |  |  |  |
| **Hospital Region** |  |  |  |  |
| Northeast | 1 |  |  |  |
| Midwest | 0.87 | 0.72 | 1.04 | 0.118 |
| South | 0.73 | 0.63 | 0.85 | <0.001 |
| West | 1.22 | 1.04 | 1.44 | 0.013 |
| **Year** |  |  |  |  |
| 2016 | 1 |  |  |  |
| 2017 | 1.06 | 0.90 | 1.26 | 0.486 |
| 2018 | 1.27 | 1.07 | 1.50 | 0.006 |
| 2019 | 1.46 | 1.24 | 1.72 | <0.001 |

Data: 2016–2019 National Inpatient Sample (NIS). aOR: adjusted Odds Ratio. CI: Confidence Interval.

^1^P value is determined using the chi-square test.

^2^The NIS defines Non-Hispanic Other as anyone who reports a race/ethnicity that does not fit in the preceding categories.

^3^Other primary payers include Worker's Compensation, CHAMPUS, CHAMPVA, Title V, and other government programs.
